# Inflammation and late-life depressive symptoms

**DOI:** 10.64898/2026.06.05.26354416

**Authors:** M. Forbes, M. Lotfaliany, B. Mengist, A-M. Isvoranu, C. Yu, P. Lacaze, M. Kang, T. Nguyen, R. L. Woods, J. J. McNeil, J. T. Neuman, M. Mohebbi, M. Berk

## Abstract

**Background:** Low-level systemic inflammation has been associated with late-life depressive symptoms. Whether individuals with higher inflammation derive preventive benefit from low-dose aspirin therapy is unknown.

**Methods:** We performed a post-hoc analysis of the ASPiring in Reducing Events in the Elderly (ASPREE) randomised, double-blind, placebo-controlled trial. Baseline C-reactive protein (hsCRP) was measured in plasma and depressive symptoms were assessed annually using the Center for Epidemiologic Studies Depression 10 Scale (CES-D-10) with elevated symptoms defined as CES-D-10 ≥ 8. Participants with elevated depressive symptoms at baseline were excluded. We fitted population-averaged logistic generalised estimating equation models adjusted for baseline sociodemographic and lifestyle covariates, including an hsCRP × treatment interaction to test effect modification by aspirin.

**Results:** Higher baseline hsCRP was associated with increased odds of elevated depressive symptoms during follow-up (OR 1.07 per SD increase in hsCRP, 95% CI 1.03–1.11). Low-dose aspirin allocation did not modify the hsCRP-depressive symptoms association (interaction OR 1.02, 95% CI 0.94–1.10).

Findings were similar after additional adjustment for comorbidity and other covariates.

**Conclusions:** In community-dwelling older adults during the ASPREE randomised trial period, higher baseline hsCRP was modestly associated with elevated depressive symptoms. There was no evidence that low-dose aspirin was associated with reduced risk of depressive symptoms among participants with higher baseline inflammation.

## Introduction

Major depression affects about 2% of community-dwelling older adults (1), however larger proportions of the population suffer from distressing depressive symptoms (2, 3). Late-life depressive symptoms emerge from a complex interplay between genetic vulnerability and environmental and biological exposures, are associated with chronic medical illnesses and impaired cognitive ability, and contribute to individual and family suffering (4). Understanding modifiable determinants of late-life depressive symptoms is a priority for healthy ageing.

Low-grade systemic inflammation may contribute to the development and persistence of depressive symptoms in older adults (5). C-reactive protein (CRP), an acute-phase protein produced by the liver in response to immune activation, has been identified as a risk biomarker (6) and predictor of later development of depression (7). This study addresses a pre-specified effect modification hypothesis, outlined in Berk et al (8), that aspirin use will reduce the risk of the onset of depressive symptoms in individuals with low-grade inflammation, as measured by high sensitivity CRP (hsCRP).

## Methods

### Study design and participants

The Aspirin for the Prevention of Depression in the Elderly (ASPREE-D) study (8) was a sub-study of the ASPirin in Reducing Events in the Elderly (ASPREE) randomised trial (9, 10). ASPREE was a double-blind, placebo-controlled trial of daily low-dose aspirin in generally healthy older adults. Participants were community-dwelling, without a history of cardiovascular disease events, were free of dementia, and had no major physical disabilities at baseline (exclusion criteria included clinical dementia or Modified Mini–Mental State Examination score <78, difficulty with or unable to independently complete six basic activities of daily living, high bleeding risk, or life-threatening illness). Between 2010 and 2014, 19,114 participants were randomised to aspirin or placebo and followed until mid-June 2017, with a median follow-up of 4.7 years.

For the present analysis, we restricted the sample to participants with available baseline hsCRP from the ASPREE Healthy Ageing Biobank (11) and at least one valid CES-D-10 assessment during follow-up. Treatment exposure was defined using intention-to-treat randomised allocation (aspirin vs placebo). Participants with elevated depressive symptoms at baseline (CES-D-10 ≥ 8) were excluded to define an at-risk cohort for longitudinal analyses. The analytic sample for each model was defined using complete-case data for variables included in that model. ASPREE was approved by ethics committees in Australia and the United States, and all participants provided written informed consent.

### Depressive symptom measurement

Depressive symptoms were measured annually using the 10-item Center for Epidemiologic Studies Depression scale (CES-D-10). The CES-D-10 is a shortened version of the original CES-D 20-item scale and has been validated for use in older adults. Scores range from 0 to 30, with higher scores indicating more depressive symptoms. We utilised a cut-score of 8 for clinically significant depressive symptoms (12, 13), as outlined in pre-specified hypotheses (8).

### C-reactive protein measurement

Non-fasting blood samples were collected at baseline and again in the third year of the trial as part of the ASPREE Biobank sub-study (11). Baseline biobank sampling was not uniformly timed relative to commencement of the trial medication. Approximately 66% of baseline samples were collected prior to starting randomised study medication and a further 14% were collected within one month of commencing study medication (11). Plasma was separated from EDTA blood samples and stored at –80°C within 4 hours of collection to preserve analytes. High sensitivity C-reactive protein (hsCRP) concentrations were measured in these stored plasma samples in a central laboratory in the Biomarker Laboratory at the University Heart & Vascular Centre, Hamburg, Germany. HsCRP was assayed using a high-sensitivity immunoturbidimetric method (an enzyme-linked assay by Abbott Diagnostics on an ARCHITECT i2000SR platform) with values reported in mg/L. The analytic platform was calibrated to detect hsCRP levels in the low range (detection limit ∼0.1 mg/L) suitable for epidemiologic use, with a coefficient of variation of < 5%. For this analysis we used the baseline hsCRP values. Because hsCRP was right-skewed, values were transformed using log (1 + hsCRP) prior to modelling.

### Covariate adjustment

Covariates were chosen to address confounding of the baseline hsCRP–depressive symptoms association while keeping the primary model parsimonious. In the primary GEE model, we adjusted for randomised treatment allocation (aspirin vs placebo) and baseline demographic and socioeconomic variables (age, sex, ethnicity, educational attainment, and living status) and included follow-up wave. In a secondary model, we additionally adjusted for alcohol and tobacco use, body mass index, dyslipidaemia, diabetes, chronic kidney disease, and polypharmacy. Models used observation-level complete-case data for the variables included, allowing participants to contribute varying numbers of follow-up observations. Effect modification by aspirin was assessed by adding a baseline hsCRP × intention-to-treat treatment interaction term to both models. We did not adjust for antidepressant use, non-steroidal anti-inflammatory drug use, or corticosteroid use because these treatments may lie on the causal pathway or induce bias through conditioning on colliders.

## Statistical analysis

Baseline characteristics were summarised at randomisation using means (SD) or median (IQR) for continuous variables and counts (%) for categorical variables. The primary outcome was elevated depressive symptoms at each follow-up visit, defined as CES-D-10 ≥ 8 and participants with baseline CES-D-10 ≥ 8 were excluded. All analyses were restricted to the randomised trial period and used intention-to-treat assignment (aspirin vs placebo). Baseline hsCRP was right-skewed so was analysed as z-standardised log (1 + hsCRP) and categorised descriptively as < 3 mg/L versus ≥ 3 mg/L.

We modelled repeated outcomes using population-averaged logistic regression fit with generalised estimating equations (GEE) with an exchangeable working correlation structure and robust standard errors. GEE was chosen in preference to time-to-event models because depressive symptoms were assessed at discrete annual visits and may fluctuate over time. A first-event approach would discard subsequent episodes and remission and could be overly sensitive to transient elevations in CES-D-10 that may represent normal and transient sadness rather than persistent depressive symptoms. Accordingly, GEE provides an interpretable estimate of the average association between baseline hsCRP and the odds of elevated depressive symptoms across follow-up.

We fitted two models: (1) a primary model adjusted for baseline sociodemographic covariates and time (visit wave), and (2) a secondary model additionally adjusted for baseline comorbidity (BMI, dyslipidaemia, diabetes, chronic kidney disease, and polypharmacy). Effect modification by aspirin was assessed by adding an hsCRP × treatment interaction term. Analyses used observation-level complete-case data for variables included in each model; follow-up visits with missing outcome or covariates were excluded, and participants could contribute varying numbers of follow-up observations.

As a sensitivity analysis, models were repeated after excluding participants with baseline hsCRP ≥ 10 mg/L, which often represents acute inflammation. Furthermore, because baseline biobank blood collection was not uniformly timed relative to commencement of trial medication, we conducted a sensitivity analysis restricted to participants whose baseline biobank sample was collected prior to starting randomised ASPREE study medication. Within this subgroup, we repeated the primary and secondary GEE models (including hsCRP × treatment interaction terms). Tests were two-sided with α = 0.05. Exploratory symptom network analyses were conducted as described in the Supplementary Methods.

## Results

### Sample characteristics

The analytic cohort comprised 10,476 participants, contributing 40,410 CES-D-10 observations over a median follow-up of 4.7 years (IQR 3.6 – 5.7 years). Participants with hsCRP ≥ 3 mg/L were slightly older, had higher BMI, were more likely to be current smokers, and had lower educational attainment (Table 1).

**Table 1.** Baseline characteristics between low and high hsCRP concentrations.

|  | hsCRP < 3 mg/L<br>(N = 7527) | hsCRP ≥ 3 mg/L<br>(N = 2949) | Total<br>(N = 10476) |
| --- | --- | --- | --- |
| Age (mean, SD) | 74.9 (4.155) | 75.3 (4.267) | 75 (4.2) |
| **Female sex (n, %)** | 3797 (50.4%) | 1700 (57.6%) | 5497 (52.5%) |
| **White ethnicity (n, %)** | 7417 (98.5%) | 2927 (99.3%) | 10344 (98.7%) |
| **Living with spouse, family or friends (n, %)** | 5314 (70.6%) | 1975 (67.0%) | 7289 (69.6%) |
| **Greater than 12 years of education (n, %)** | 4024 (53.5%) | 1380 (46.8%) | 5404 (51.6%) |
| **Current alcohol use (n, %)** | 6100 (81.0%) | 2298 (77.9%) | 8398 (80.2%) |
| **Current smoking (n, %)** | 185 (2.5%) | 126 (4.3%) | 311 (3.0%) |
| **Body mass index\* (mean, SD)** | 27.3 (4.1) | 29.6 (4.9) | 27.9 (4.4) |
| **Dyslipidaemia\*<sup>†</sup> (n, %)** | 5332 (72.6%) | 2230 (77.0%) | 7562 (73.8%) |
| **Chronic kidney disease\*<sup>†</sup> (n, %)** | 1174 (16.1%) | 602 (21.0%) | 1776 (17.5%) |
| **Diabetes\*<sup>†</sup> (n, %)** | 541 (7.3%) | 279 (9.6%) | 820 (8.0%) |
| **Polypharmacy<sup>†</sup> (n, %)** | 2095 (27.8%) | 1092 (37.0%) | 3187 (30.4%) |
| **Antidepressant use<sup>‡</sup> (n, %)** | 713 (9.5%) | 369 (12.5%) | 1082 (10.3%) |
| **hsCRP (median, IQR)** | 1.2 (0.71–1.84) | 5.12 (3.79–8.03) | 1.67 (0.88–3.31) |
| **CES-D-10 score (median, IQR)** | 2 (0–4) | 2 (1–4) | 2 (0–4) |

\* Missing values were BMI n=43 (0.4%; 29 in hsCRP <3 mg/L and 14 in hsCRP ≥3 mg/L), dyslipidaemia n=231 (2.2%; 179 and 52), chronic kidney disease n=306 (2.9%; 220 and 86), and diabetes n=195 (1.9%; 139 and 56). Percentages were calculated using non-missing denominators.
<sup>†</sup> Dyslipidaemia was defined as lipid-lowering medication use and/or elevated total cholesterol (≥ 212 mg/dL [Australia] or ≥ 240 mg/dL [USA]). Diabetes was defined as self-reported diabetes, glucose-lowering treatment, and/or fasting glucose ≥ 126 mg/dL (7.0 mmol/L). Chronic kidney disease was defined as eGFR < 60 mL/min/1.73 m<sup>2</sup> and/or UACR ≥ 3 mg/mmol. Polypharmacy was defined as use of ≥ 5 regular prescription medications at baseline.
<sup>‡</sup> Antidepressant use was defined as use of any N06A (ATC/DDD index) medications at baseline.

### Association between baseline hsCRP and elevated depressive symptoms during follow-up

In the primary GEE model (n = 10,476; 40,410 observations), higher baseline hsCRP was associated with increased odds of elevated depressive symptoms during follow-up (OR 1.07 per SD, 95% CI 1.03–1.11) (Table 2). Randomised aspirin allocation was not associated with elevated depressive symptoms (OR 0.96, 95% CI 0.89–1.04), and there was no evidence that aspirin modified the hsCRP association (hsCRP × treatment OR 1.02, 95% CI 0.94–1.10). In the second model, adjusted for alcohol and tobacco use, BMI, dyslipidaemia, diabetes, chronic kidney disease and polypharmacy (n = 10,192; 32,187 observations), the hsCRP association was attenuated (OR 1.04, 95% CI 1.00–1.09). Again, there was no evidence of effect modification (hsCRP × treatment OR 1.01, 95% CI 0.93–1.10).

**Table 2.** Association of baseline hsCRP with elevated depressive symptoms during follow-up and test of modification by randomised aspirin allocation

|  | Model 1 OR<br>(95% CI) | Model 1 +<br>interaction OR<br>(95% CI) | Model 2 OR<br>(95% CI) | Model 2 +<br>interaction OR<br>(95% CI) |
| --- | --- | --- | --- | --- |
| **Baseline hsCRP (per SD)** | 1.07 (1.03–1.11) | 1.06 (1.01–1.12)<sup>†</sup> | 1.04 (1.00–1.09) | 1.03 (0.97–1.10)<sup>†</sup> |
| **Aspirin vs placebo** | 0.96 (0.89–1.04) | 0.96 (0.89–1.04)<sup>†</sup> | 0.98 (0.90–1.07) | 0.98 (0.90–1.07)<sup>†</sup> |
| **hsCRP × Aspirin** | — | 1.02 (0.94–1.10) | — | 1.01 (0.93–1.10) |
| **Participants (n)\*** | 10,476 | 10,476 | 10,192 | 10,192 |
| **Observations** | 40,410 | 40,410 | 32,187 | 32,187 |

Model 1 was adjusted for age, sex, ethnicity, living status and educational level. Model 2 was adjusted for age, sex, ethnicity, living status, educational level, alcohol use, tobacco use, body mass index, dyslipidaemia, diabetes, chronic kidney disease and polypharmacy
<sup>†</sup> In interaction models, hsCRP is the effect in the placebo group; the interaction term indicates if this differs under aspirin.
\* Participants denotes unique individuals contributing ≥ 1 complete follow-up observation; Observations denotes complete follow-up visits after observation-level complete-case exclusion. Model 2 additionally included adjustment for alcohol, smoking, BMI, dyslipidaemia, diabetes, CKD, and polypharmacy

Figure 1 presents the observed (unadjusted) prevalence of elevated depressive symptoms across follow-up by baseline hsCRP group and treatment allocation. It demonstrates that within hsCRP strata, aspirin and placebo trajectories were similar with overlapping confidence intervals.

**Figure 1.**
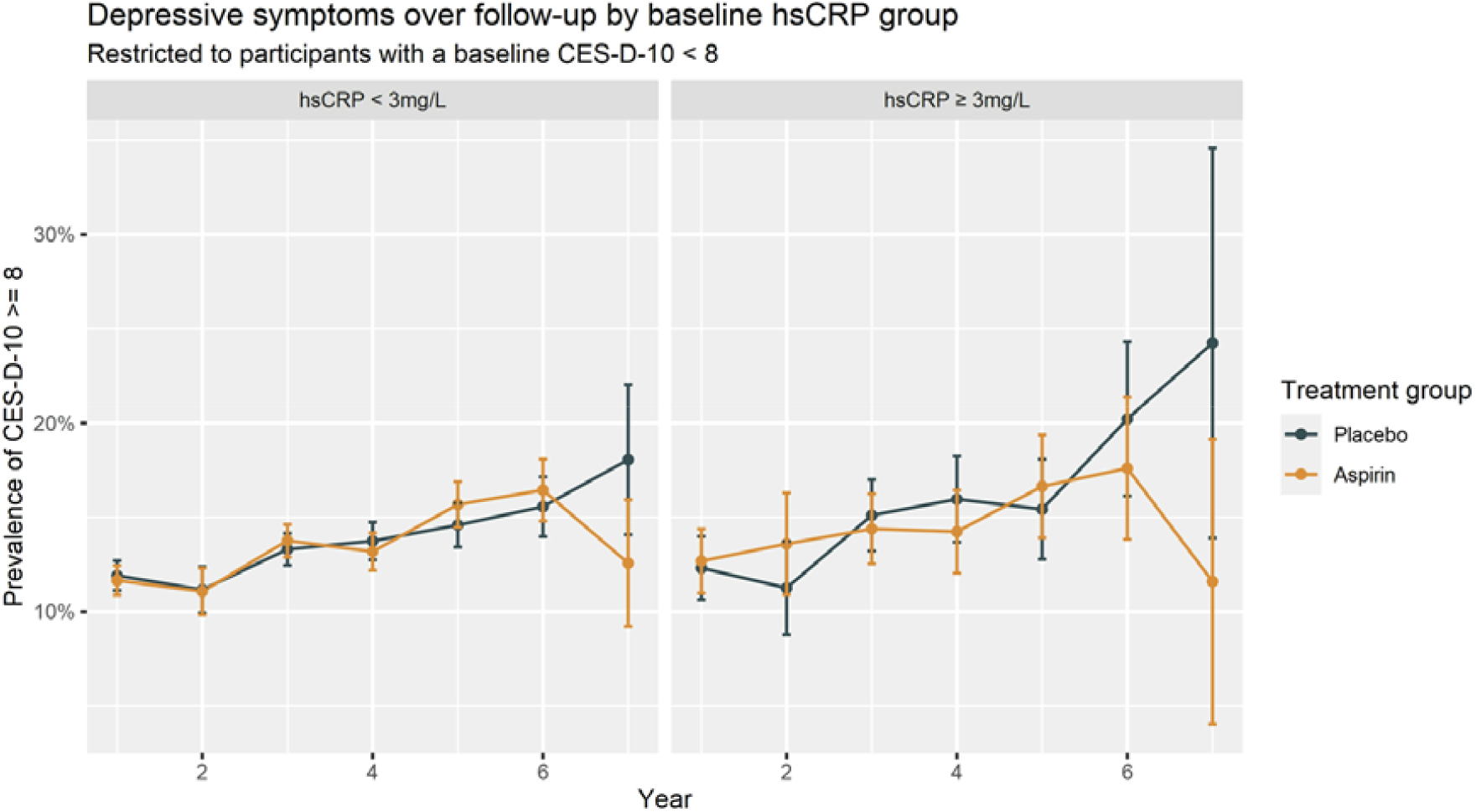
Observed prevalence of elevated depressive symptoms over follow-up by baseline hsCRP group and randomised treatment allocation (aspirin vs placebo)

### Sensitivity analyses

After excluding participants with baseline hsCRP ≥ 10 mg/L, which is more likely to reflect acute inflammation, results were broadly consistent. In the primary model (n = 9,953; 38,437 observations), hsCRP was associated with elevated depressive symptoms (OR 1.10, 95% CI 1.05– 1.16). In the second model, adjusted for clinical variables (n = 9,696; 30,646 observations), the association remained (OR 1.08, 95% CI 1.02–1.14). There was no evidence of effect modification by aspirin in either model.

In analyses restricted to participants whose baseline biobank sample was collected prior to starting trial medication (n = 6,947; 26,794 observations for Model 1), the primary hsCRP association persisted (OR 1.07, 95% CI 1.02–1.12) and there was no evidence of hsCRP × aspirin interaction. In the fully adjusted model (n = 6,773; 21,433 observations), the hsCRP association attenuated (OR 1.04, 95% CI 0.98–1.10) and the interaction remained non-significant (Table S1).

## Discussion

In this prospective analysis of community-dwelling older adults without elevated depressive symptoms at baseline, higher baseline hsCRP was associated with a modest increase in the odds of elevated depressive symptoms during follow-up. However, this association attenuated after adjustment for baseline comorbidity, suggesting that part of the CRP-depressive symptoms relationship may reflect underlying poor physical health. Across models, we found no evidence that low-dose aspirin therapy modified the association between hsCRP and elevated depressive symptoms.

The observed effect size for hsCRP was small, consistent with systemic inflammation being one contributor among many to late-life depressive symptoms. The attenuation after adjustment for comorbidity accords with prior findings that associations between hsCRP and depressive symptoms in adults can be partly explained by poor physical health and cardiometabolic disease (14, 15). In this context, hsCRP may operate as a non-specific marker of adverse ageing processes (e.g., cardiometabolic risk, subclinical disease, frailty-related physiology). However, large observational studies suggest the association between systemic inflammation and depressive symptoms persists even after extensive adjustment for physical health (16).

Assuming that C-reactive protein does have an independent role in the pathogenesis of depressive symptoms, how can we explain the null effect of low-dose aspirin? A plausible explanation is that low-dose aspirin does not significantly reduce low-grade systemic inflammation in a relatively healthy older cohort. Several trials have reported no meaningful reduction in CRP with aspirin (17, 18). It remains possible that other anti-inflammatory agents with greater immunomodulatory effects may exert antidepressant effects in selected high-inflammation subgroups, and the Advanced Stratification of People with Depression Based on Inflammation (ASPIRE) study is underway to address this question (19).

Inflammation may also relate more strongly to specific symptom domains than to depression as a unitary construct. Prior symptom-level work links inflammatory markers most consistently to neurovegetative or somatic features (e.g., low energy, fatigue, sleep disturbance, and appetite changes) (20, 21). Our exploratory symptom network analyses were consistent with this pattern, with hsCRP showing conditional associations with items reflecting effort/fatigue and reduced drive, aligning with “sickness behaviour” models (22).

Strengths of this study include the large, well-characterised ASPREE cohort, repeated standardised assessment of depressive symptoms, biomarker measurement from the ASPREE biobank, and the ability to test a prespecified effect-modification hypothesis within a randomised trial framework using intention-to-treat allocation. Limitations include the use of annual symptom assessment with a screening scale rather than diagnostic interviews, and hsCRP measured at baseline (and year 3 for the secondary biomarker analysis) rather than repeatedly at annual visits. Baseline hsCRP was derived from biobanked samples and collection timing was not uniformly pre-randomisation, although sensitivity analyses restricted to participants whose baseline sample was collected prior to commencing trial medication produced similar conclusions. HsCRP is also a non-specific marker and may be influenced by intercurrent illnesses or clinical events around blood collection, potentially adding noise. Although we adjusted for major comorbidity and medication burden, comorbidities were primarily based on baseline participant report and residual confounding cannot be excluded. The cohort was predominantly White and selected for good baseline health, which may limit generalisability to more diverse or medically complex older populations. Finally, the fully adjusted models used complete-case analysis. While missingness was low, some selection bias is possible.

## Conclusion

In this large cohort of community-dwelling older adults followed prospectively during the ASPREE randomised trial, higher baseline hsCRP was associated with a modest elevation in the odds of later depressive symptoms. This association was attenuated after adjustment for lifestyle factors and comorbidity. Low-dose aspirin did not modify the hsCRP-depressive symptoms association. Together, these findings do not support using aspirin as a preventive strategy for depressive symptoms among older adults with elevated hsCRP. Future research is warranted to investigate inflammatory phenotyping and symptom-domain profiling and to test interventions with clearer immunomodulatory effects to determine whether any subgroups derive clinically meaningful benefit from inflammation-targeted interventions.

## Data Availability

Data are available on reasonable request.

https://ams.aspree.org/public/

## Supplement A

### Methods

We conducted an exploratory, cross-sectional symptom network analysis to examine whether systemic inflammation, as measured by hsCRP, was preferentially related to specific depressive symptom. At baseline and year 3, we estimated regularised partial correlation networks including log-transformed hsCRP and the 10 CES-D-10 items as nodes using established methods (23, 24).

The network included 11 nodes: log (1 + CRP) and the 10 CES-D items scored on the original 0–3 response scale (higher scores indicating greater symptom burden). CRP was measured in mg/L and transformed as log(1+CRP) to reduce right-skew and avoid undefined values at CRP = 0. Items were: Cesd1 “Bothered by things”, Cesd2 “Trouble keeping mind on task”, Cesd3 “Felt depressed”, Cesd4 “Everything was an effort”, Cesd5 “Felt less hopeful”, Cesd6 “Felt fearful”, Cesd7 “Restless sleep”, Cesd8 “Felt less happy”, Cesd9 “Felt lonely”, and Cesd10 “Couldn’t get going”.

Networks were estimated separately at baseline and year 3 using EBICglasso (graphical LASSO), yielding sparse undirected networks in which edges represent partial correlations between nodes, conditioned on all other nodes. Spearman correlations were used to reduce sensitivity to non-normality and the ordinal nature of symptom responses. The EBIC hyperparameter was set to γ=0.5. Baseline and year 3 networks were visualised using a shared node layout to facilitate comparison; edge colour indicated sign and edge thickness reflected magnitude.

At each wave, analyses were restricted to participants with complete data on all 11 nodes (CRP and all 10 CES-D items). For the paired baseline-year 3 comparison, analyses were restricted to participants with complete node data at both waves.

Edge-weight accuracy was evaluated using nonparametric bootstrap procedures (95% confidence intervals). Centrality indices were computed descriptively, with primary emphasis on strength. Strength stability was assessed using case-dropping procedures and summarised by the correlation stability (CS) coefficient. Baseline and year 3 networks were compared using the paired Network Comparison Test (NCT) to test differences in (i) network structure and (ii) global strength. Individual edge differences were examined exploratorily with Benjamini–Hochberg adjustment. All network findings were interpreted descriptively; networks are undirected and do not imply causality or directionality.

### Results

The baseline network was estimated in 11,942 participants with complete node data, and the year 3 network in 10,339 participants. The paired baseline–year 3 NCT comparison included 10,153 participants with complete node data at both time points.

Supplementary Figures S1 and S2 show the estimated EBICglasso networks at baseline and year 3, respectively. In both waves, CRP showed its most prominent retained conditional associations with “everything was an effort” (Cesd4) and “couldn’t get going” (Cesd10), consistent with a neurovegetative/somatic symptom pattern. The CRP–Cesd4 regularised partial correlation was 0.0406 at baseline and 0.0417 at year 3; the CRP–Cesd10 regularised partial correlation was 0.0298 at baseline and 0.0433 at year 3.

**Figure S1:**
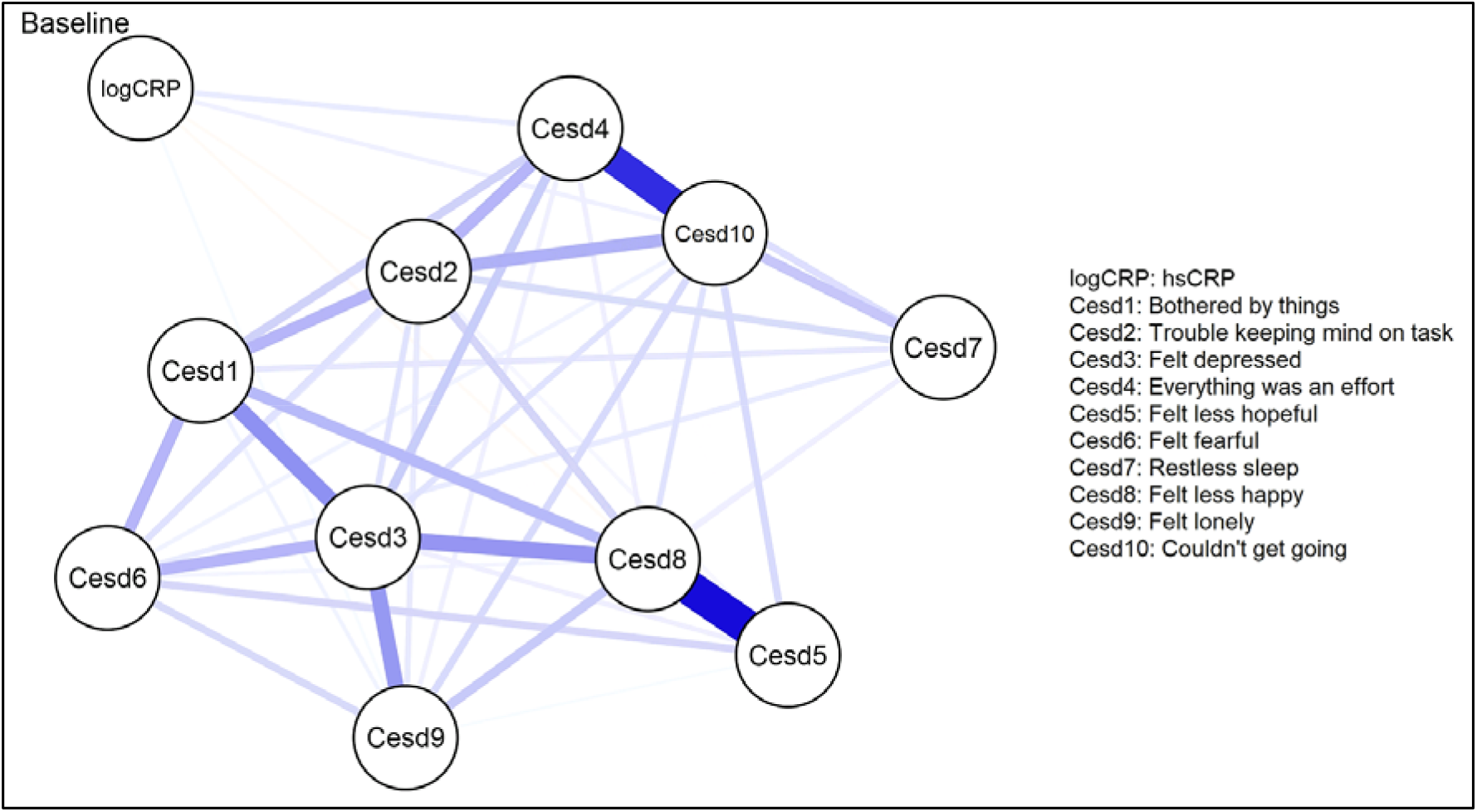
Regularised partial correlation networks at baseline (edge thickness reflects magnitude and colour indicates sign with blue being a positive association)

**Figure S2:**
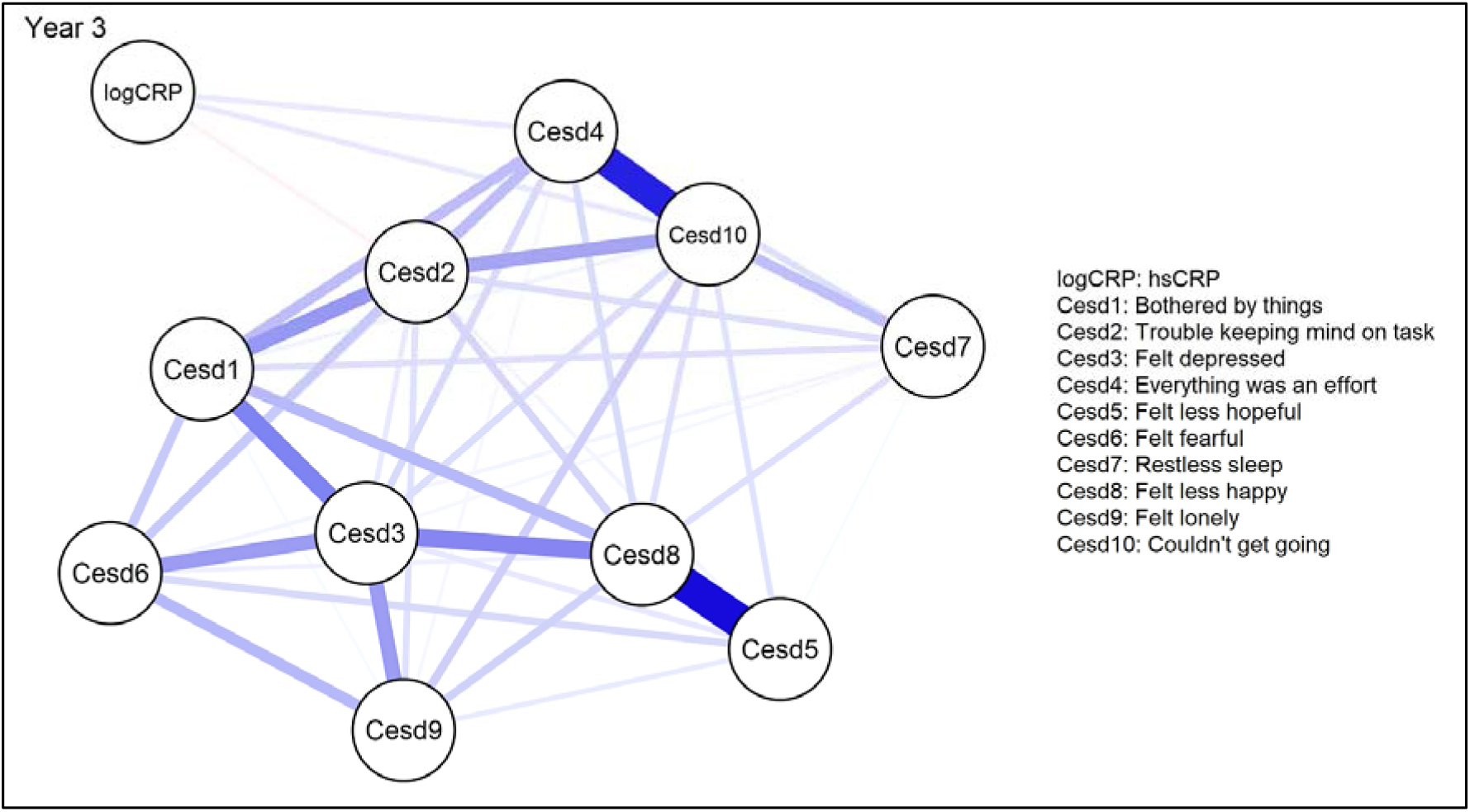
Regularised partial correlation networks at year 3 (edge thickness reflects magnitude and colour indicates sign with blue being a positive association)

Bootstrap edge-weight accuracy plots are shown in Supplementary Figures S3 and S4.Strength centrality stability was high in both networks (CS = 0.75 at baseline and CS = 0.75 at year 3), indicating stable strength estimates under case-dropping. The paired NCT provided limited evidence for a difference in overall network structure between baseline and year 3 (M = 0.0567, p = 0.088). In contrast, global strength was higher at year 3 than baseline (3.8737 vs 3.4607; S = 0.4130, p = 0.0005), suggesting increased overall connectivity among nodes at year 3. Edges estimated as exactly zero reflect regularisation under EBICglasso and should be interpreted as “no retained conditional association under the selected penalty” rather than definitive evidence of absence.

**Figure S3:**
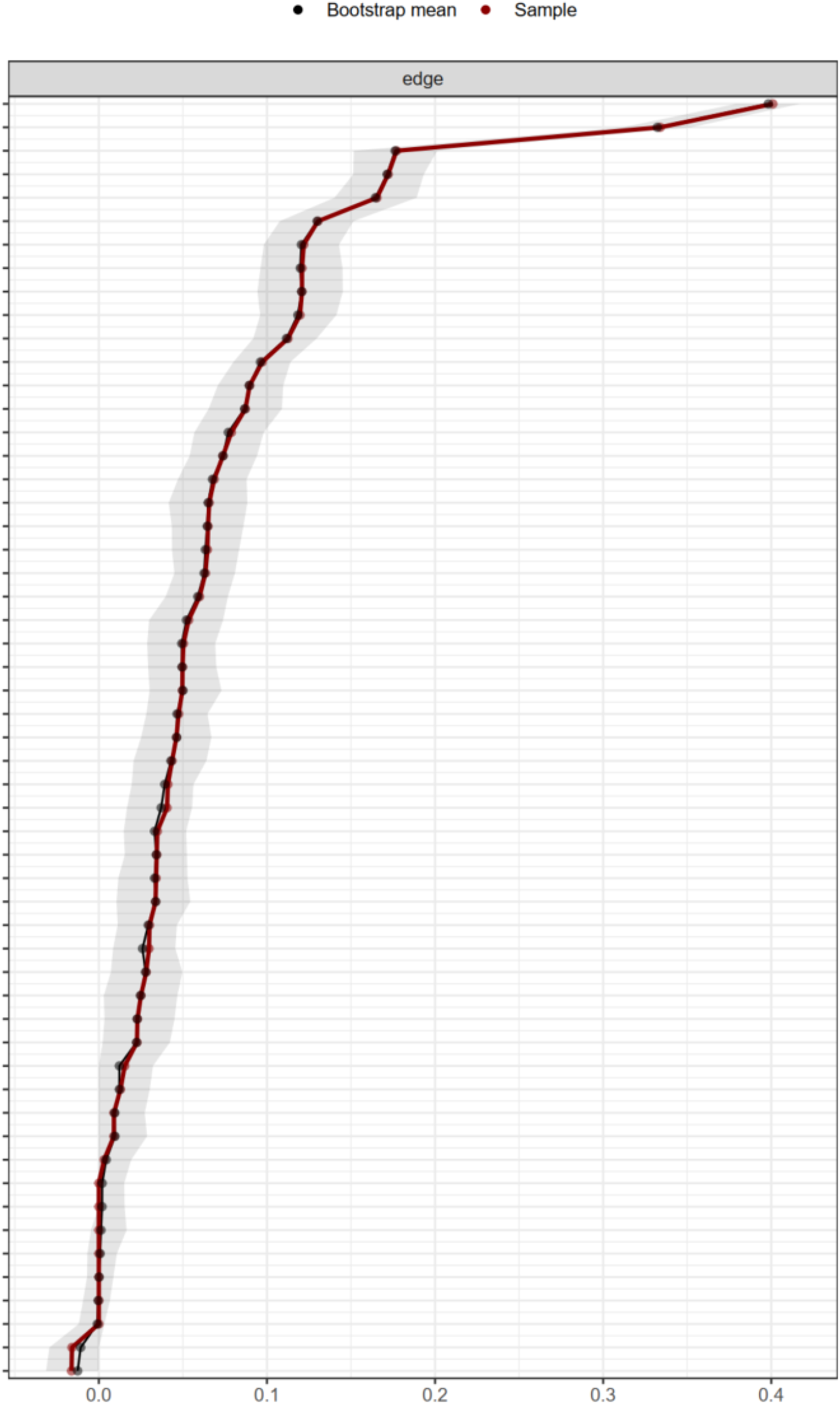
Bootstrap 95% confidence intervals for baseline network edge weights (nonparametric bootstrap)

**Figure S4:**
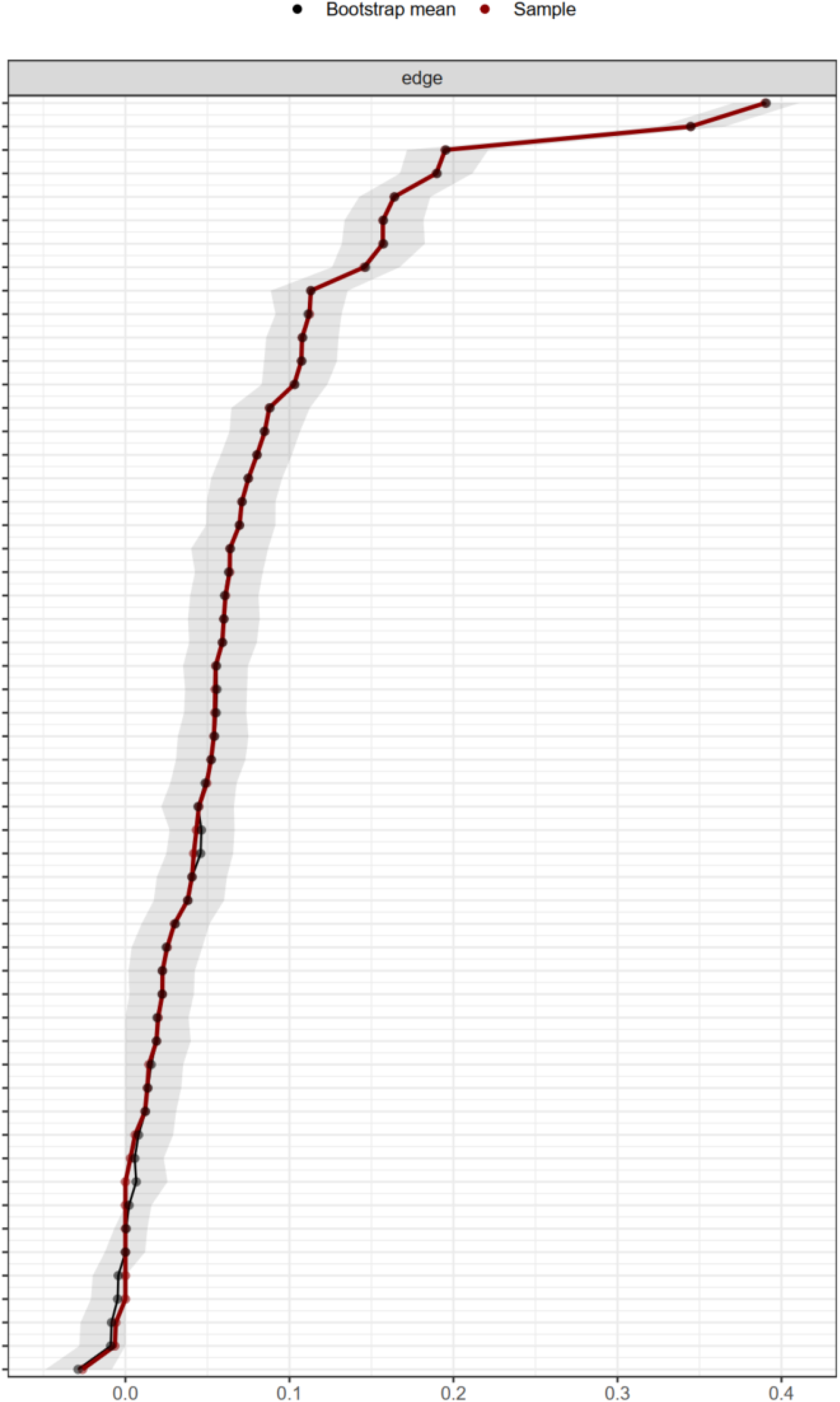
Bootstrap 95% confidence intervals for year 3 network edge weights (nonparametric bootstrap)

## Supplement B

**Table S1:**
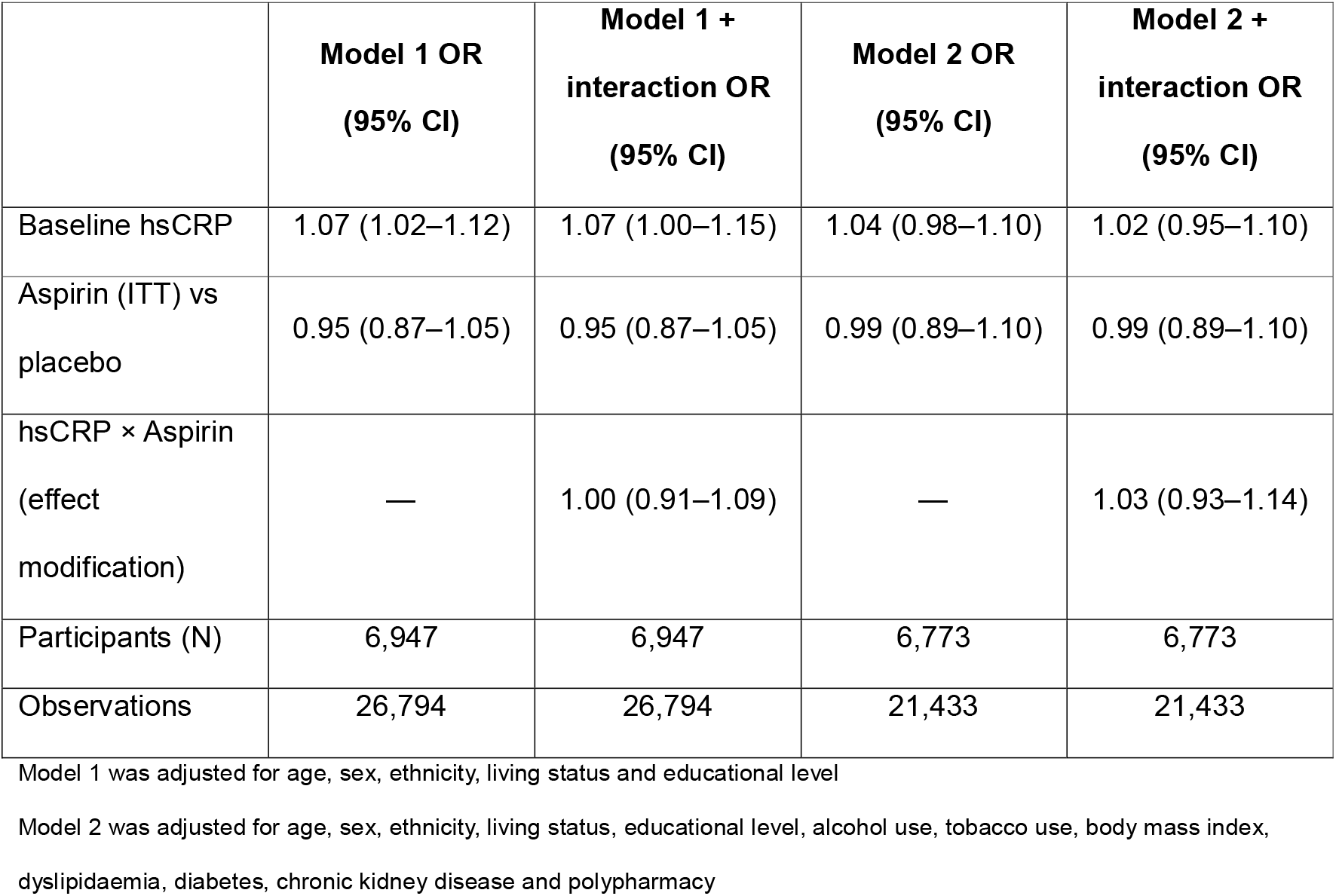
Association of baseline hsCRP with elevated depressive symptoms during follow-up and test of modification by aspirin allocation, restricted to participants whose baseline biobank sample was collected prior to starting trial medication

## Acknowledgements

The authors express their gratitude to the participants and staff involved in data collection and management of ASPREE.

## References

1. Beekman AT, Copeland JR, Prince MJ. Review of community prevalence of depression in later life. Br J Psychiatry. 1999;174:307–11.

2. Buchtemann D, Luppa M, Bramesfeld A, Riedel-Heller S. Incidence of late-life depression: a systematic review. J Affect Disord. 2012;142(1-3):172–9.

3. Andreas S, Schulz H, Volkert J, Dehoust M, Sehner S, Suling A, et al. Prevalence of mental disorders in elderly people: the European MentDis_ICF65+ study. Br J Psychiatry. 2017;210(2):125–31.

4. Alexopoulos GS. Depression in the elderly. Lancet. 2005;365(9475):1961–70.

5. Mac Giollabhui N, Ng TH, Ellman LM, Alloy LB. The longitudinal associations of inflammatory biomarkers and depression revisited: systematic review, meta-analysis, and meta-regression. Mol Psychiatry. 2021;26(7):3302–14.

6. Zeng Y, Chourpiliadis C, Hammar N, Seitz C, Valdimarsdóttir UA, Fang F, et al. Inflammatory Biomarkers and Risk of Psychiatric Disorders. JAMA Psychiatry. 2024;81(11):1118–29.

7. Pasco JA, Nicholson GC, Williams LJ, Jacka FN, Henry MJ, Kotowicz MA, et al. Association of high-sensitivity C-reactive protein with de novo major depression. Br J Psychiatry. 2010;197(5):372–7.

8. Berk M, Woods RL, Nelson MR, Shah RC, Reid CM, Storey E, et al. ASPREE-D: Aspirin for the prevention of depression in the elderly. Int Psychogeriatr. 2016;28(10):1741–8.

9. ASPREE Investigator Group. Study design of ASPirin in Reducing Events in the Elderly (ASPREE): a randomized, controlled trial. Contemp Clin Trials. 2013;36(2):555–64.

10. McNeil JJ, Woods RL, Nelson MR, Reid CM, Kirpach B, Wolfe R, et al. Effect of Aspirin on Disability-free Survival in the Healthy Elderly. N Engl J Med. 2018;379(16):1499–508.

11. Parker EJ, Orchard SG, Gilbert TJ, Phung JJ, Owen AJ, Lockett T, et al. The ASPREE Healthy Ageing Biobank: Methodology and participant characteristics. PLoS One. 2024;19(2):e0294743.

12. Andresen EM, Malmgren JA, Carter WB, Patrick DL. Screening for Depression in Well Older Adults: Evaluation of a Short Form of the CES-D. American Journal of Preventive Medicine. 1994;10(2):77–84.

13. Mohebbi M, Agustini B, Woods RL, McNeil JJ, Nelson MR, Shah RC, et al. Prevalence of depressive symptoms and its associated factors among healthy community-dwelling older adults living in Australia and the United States. Int J Geriatr Psychiatry. 2019;34(8):1208–16.

14. Almeida OP, Norman P, Hankey GJ, Jamrozik K, Flicker L. The association between C-reactive protein concentration and depression in later life is due to poor physical health: results from the Health in Men Study (HIMS). Psychol Med. 2007;37(12):1775–86.

15. Fried EI, von Stockert S, Haslbeck JMB, Lamers F, Schoevers RA, Penninx BWJH. Using network analysis to examine links between individual depressive symptoms, inflammatory markers, and covariates. Psychological Medicine. 2020;50(16):2682–90.

16. Pitharouli MC, Hagenaars SP, Glanville KP, Coleman JRI, Hotopf M, Lewis CM, et al. Elevated C-Reactive Protein in Patients With Depression, Independent of Genetic, Health, and Psychosocial Factors: Results From the UK Biobank. Am J Psychiatry. 2021;178(6):522–9.

17. Feldman M, Jialal I, Devaraj S, Cryer B. Effects of low-dose aspirin on serum C-reactive protein and thromboxane B2 concentrations: a placebo-controlled study using a highly sensitive C-reactive protein assay. Journal of the American College of Cardiology. 2001;37(8):2036–41.

18. Kim MA, Kim CJ, Seo JB, Chung WY, Kim SH, Zo JH, et al. The effect of aspirin on C-reactive protein in hypertensive patients. Clin Exp Hypertens. 2011;33(1):47–52.

19. Worrell C, Baune BT, Benedetti F, Cattaneo A, De Picker L, Felger JC, et al. Anti-inflammatories for depression: challenges and ASPIRations. Lancet Psychiatry. 2025.

20. Duivis HE, de Jonge P, Penninx BW, Na BY, Cohen BE, Whooley MA. Depressive symptoms, health behaviors, and subsequent inflammation in patients with coronary heart disease: prospective findings from the heart and soul study. Am J Psychiatry. 2011;168(9):913–20.

21. Jokela M, Virtanen M, Batty GD, Kivimäki M. Inflammation and Specific Symptoms of Depression. JAMA Psychiatry. 2016;73(1):87–8.

22. Capuron L, Miller AH. Immune system to brain signaling: neuropsychopharmacological implications. Pharmacol Ther. 2011;130(2):226–38.

23. Burger J, Isvoranu AM, Lunansky G, Haslbeck JMB, Epskamp S, Hoekstra RHA, et al. Reporting standards for psychological network analyses in cross-sectional data. Psychol Methods. 2023;28(4):806–24.

24. Isvoranu A-M, Epskamp S, Waldorp LJ, Borsboom D. Network psychometrics with R: A guide for behavioral and social scientists. New York, NY, US: Routledge; 2022. 250–p.

